# Human Behaviour and Perceptions as Drivers for Persistent Malaria Transmission in Mudzi District, Zimbabwe: Results from Focus Group Discussions

**DOI:** 10.1101/2025.04.30.25326649

**Authors:** Tichaona Fambirai, Moses John Chimbari, Tawanda Manyangadze

## Abstract

**Objectives:** This study aims to establish knowledge, perceptions, attitudes, and practices on malaria in Mudzi district, Zimbabwe, where there is persistent malaria transmission despite high indoor residual spraying coverage.

**Design:** A qualitative study employing focused group discussions (FGDs) was used to establish knowledge, perceptions, and practices on malaria among the local population. NVIVO version 13 was used for content analysis.

**Setting:** The study was conducted in a rural district located on the Zimbabwe and Mozambique border region.

**Participants:** Participants were residents of six (6) randomly selected villages in Mudzi district, Zimbabwe, aged 15 years and above.

**Results:** Eighteen (18) FGDs were conducted with 148 individuals participating. The majority of participants were unemployed, 102 (69.9%), and had attained secondary level education, 138(94.5%). Participants showed good knowledge of malaria, but misconceptions were also reported. Participants reported high community participation in night outdoor activities, prolonged outdoor religious activities, and artisanal gold mining with minimal malaria preventative measures. Long queues at health facilities, negative attitudes of the healthcare staff, inadequate community health workers (CHWs) coverage, malaria diagnostics, and medicine stockouts were reported by participants as key hindrances to accessing malaria treatment. Indoor residual spraying (IRS) was accepted by participants as an effective malaria control intervention; however, some participants viewed IRS chemicals currently in use as less effective compared to previous ones. This perception may likely result in low IRS uptake; hence, effective education is required by program managers and spray operators.

**Conclusion:** The study revealed complex intertwined human, economic, and social behavioural practices and perceptions that minimise the protective effect of IRS. Intensifying integrated targeted social behaviour change intervention towards key groups may reduce night outdoor activities, improve risk perception, and increase uptake of personal protective measures. Critical investment should be made in expanding human resources for health and reducing access to care bottlenecks.

**Strengths and limitations of this study:**

- This study involved focus group discussions separated by gender and age. This allowed full expression of views from all groups without hindrance from cultural and social impediments.
- Data collection was done in multiple sites of the districts. This ensured that representatives and may improve the generalisability of the results.
- The study was carried out in two majorly rural districts, and results may not be generalizable to different settings.
- The selection of knowledge-rich individuals introduced bias as willing, hesitant individuals who could have been knowledgeable may have been excluded, thus depriving the FGDs of diverse views.

## Introduction

Malaria is one of the single biggest public health threats, responsible for over 230 million cases and 597,000 deaths ^[1]^. Sub-Saharan Africa contributes at least 94% of cases of the global burden ^[1]^. Significant gains have been recorded in malaria control in the last two decades. The interaction of humans and vectors is considered to be the major driver of malaria transmission, as staying outdoors for long hours at night has been associated with higher cases of malaria ^[2]^. Human outdoor movement, quality of housing, occupation, climatic factors, agricultural practices, and land use changes are the primary determinants for malaria transmission ^[3–5]^. There has been a significant reduction in the global malaria burden as a result of vector control, malaria diagnostics, malaria treatment, and surveillance. However, emerging artemisinin resistance, climate change, and diminishing global and domestic finances are threatening to reverse the progress made ^[6]^. Increased human global movement from endemic and non-endemic regions and cross-border movement has potentiated transboundary malaria parasite movement ^[7–9]^.

An association between poverty and malaria occurrence ^[10–12]^ has been established, with regions characterised by low income per capita associated with high endemicity ^[5,13]^. Poor housing and limited access to care and information have also been identified as significant drivers of malaria transmission^[14,15]^. Outdoor occupations like agriculture, mining, fishing, and forestry sectors, which involve prolonged night outdoor activities, have also been implicated as a significant determinant for contracting malaria globally^[16–18]^.

Individual and community practices, perceptions, and attitudes have also been shown to be key determinants of uptake of interventions, access to care, and predictors for adherence ^[19]^. Limited knowledge and misinterpretation influence how individuals adopt interventions and adhere to treatment ^[20]^. Increased knowledge of malaria transmission also influences the uptake and utilization of malaria-preventative measures ^[21]^. Ideation models on behaviour have revealed that humans will not act until they have enough knowledge about action and consequences and have developed a positive attitude about certain courses of action ^[22,23]^. Despite huge investments in health education, misconceptions still persist in some communities, as reported in a study by Zerdo et al. in Ethiopia ^[24]^, where communities attributed malaria infections to hunger and eating certain foods. Additionally, some communities erroneously believe they can contract malaria from drinking dirty water and due to other supernatural phenomena ^[24]^.

Malaria transmission in Zimbabwe is characterized by seasonal variations, with the wet season (November-March) associated with high transmission ^[25,26]^. The transmission is heterogeneous with low altitude and hot regions characterized by high transmission compared to the highlands ^[27]^. Significant success in reducing malaria burden has been recorded since 2003, with incidence declining by 84% ^[28,29]^.

Mudzi district experiences persistent malaria infections with an annual incidence rate >200 cases per 1000 population. The district accounts for at least 33% of the national malaria burden ^[30]^. Highly persistent malaria infections exist in the district despite high annual district IRS coverages exceeding 95%, room and population coverages. We believe that the high persistence of malaria transmission despite effective IRS coverage could be due to human behavioral factors that have not been fully studied. Thus, our study focused on these aspects and which are critical in the formulation of innovative socio-behavioural evidence-based interventions for malaria control.

## Methods

### Study Setting

Mudzi [Geo-coordinates: 17°00ʹS032°40ʹE] is one of the nine administrative districts in Mashonaland East Province, located on the eastern border region of Zimbabwe and Mozambique **Fig. 1**.

**Figure 1.**
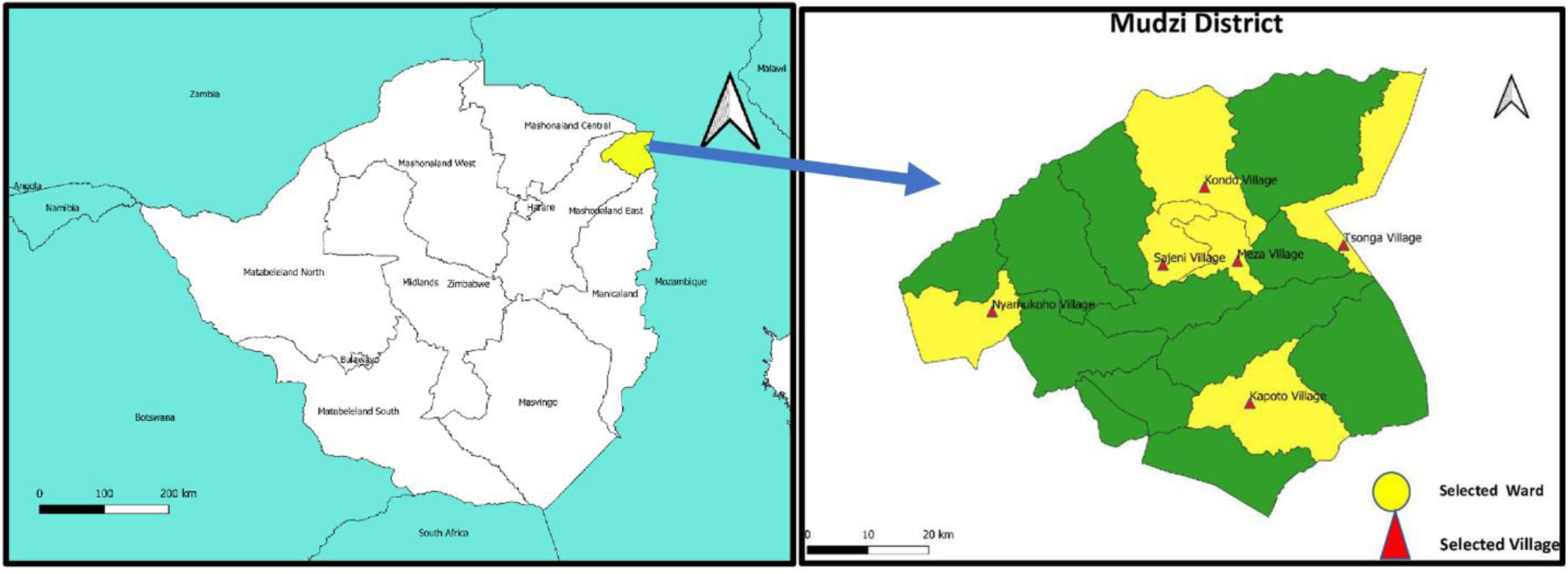
Map of Zimbabwe and Mudzi District showing selected wards and villages study sites

The local human population is estimated at 165,266 ^[31]^ based on the 2022 national census data. The district has eighteen (18) administrative wards and 523 villages. A village is a small administrative geographical unit comprising 10-50 households supervised by a village head. The district lies in the low, dry region of the country that experiences low annual rainfall and high temperatures. Due to rich alluvial gold deposits, the local population engages in artisanal gold mining (AGM) in Makaha, Rwenya River, and Mazoe River areas. A single ward (comprising 6-7 villages) is served by a single rural health center (RHC) manned by a registered general nurse (RGN), a Primary care nurse (PCN), and an Environmental Health Technician (EHT). Community Health Workers (CHW) provide community-based malaria testing and treatment services. Cross-border movement between Mudzi and Changara district (Mozambique) is common as the local population shares common historical social ties.

### Sampling

A multi-stage sampling technique was used to select study sites **(Supplementary File 1).** Eighteen (18) wards were grouped into six (6) clusters, comprised of three (3) wards. A single ward was selected from each of the clusters, resulting in the selection of six (6), villages (Tsonga, Meza, Kondo, Sajeni, Nyamukoho and Kapoto) as shown in **Fig. 2**. Villages in the selected wards were listed in Excel, and the RANDOM function was used to select a single village from each ward. A total of six villages were enrolled in the study (Tsonga, Meza, Sajeni, Magohoto, Kapoto, and Nyamukoho). Participants for each of the FGDs were recruited from each of the selected villages. Adult males (AM), adult females (AF), and a mixed group (MG) of young adults aged 16-21 were recruited to participate in the FGDs. The separation of the groups by gender and age was to allow equitable participation of community sub-groups and reduce socio-cultural impediments to participation. The local CHW assisted the researchers in recruiting study participants.

**Figure 2.**
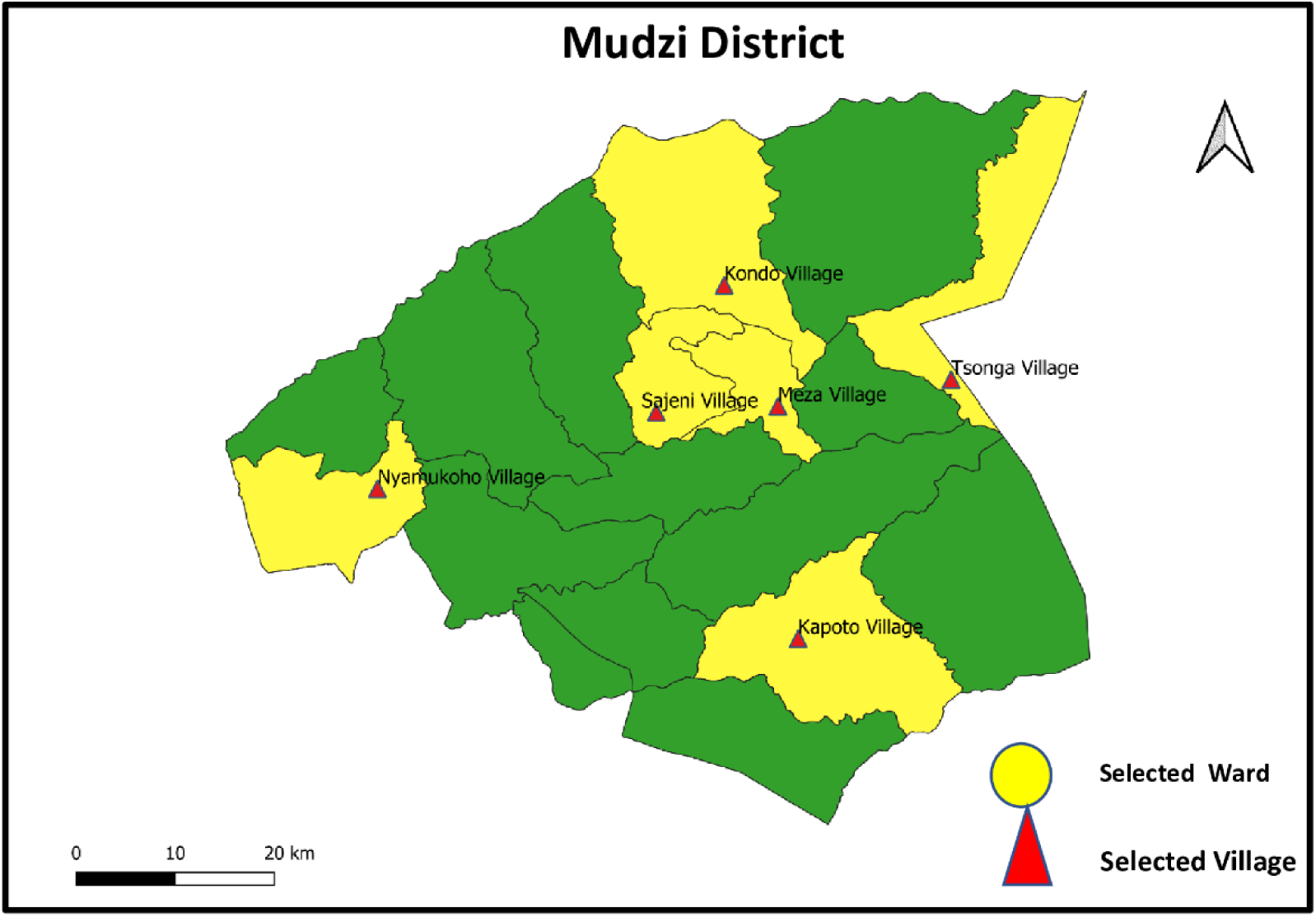
Selected Study Ward and Villages in Mudzi District, Zimbabwe

**Supplementary File 1.**
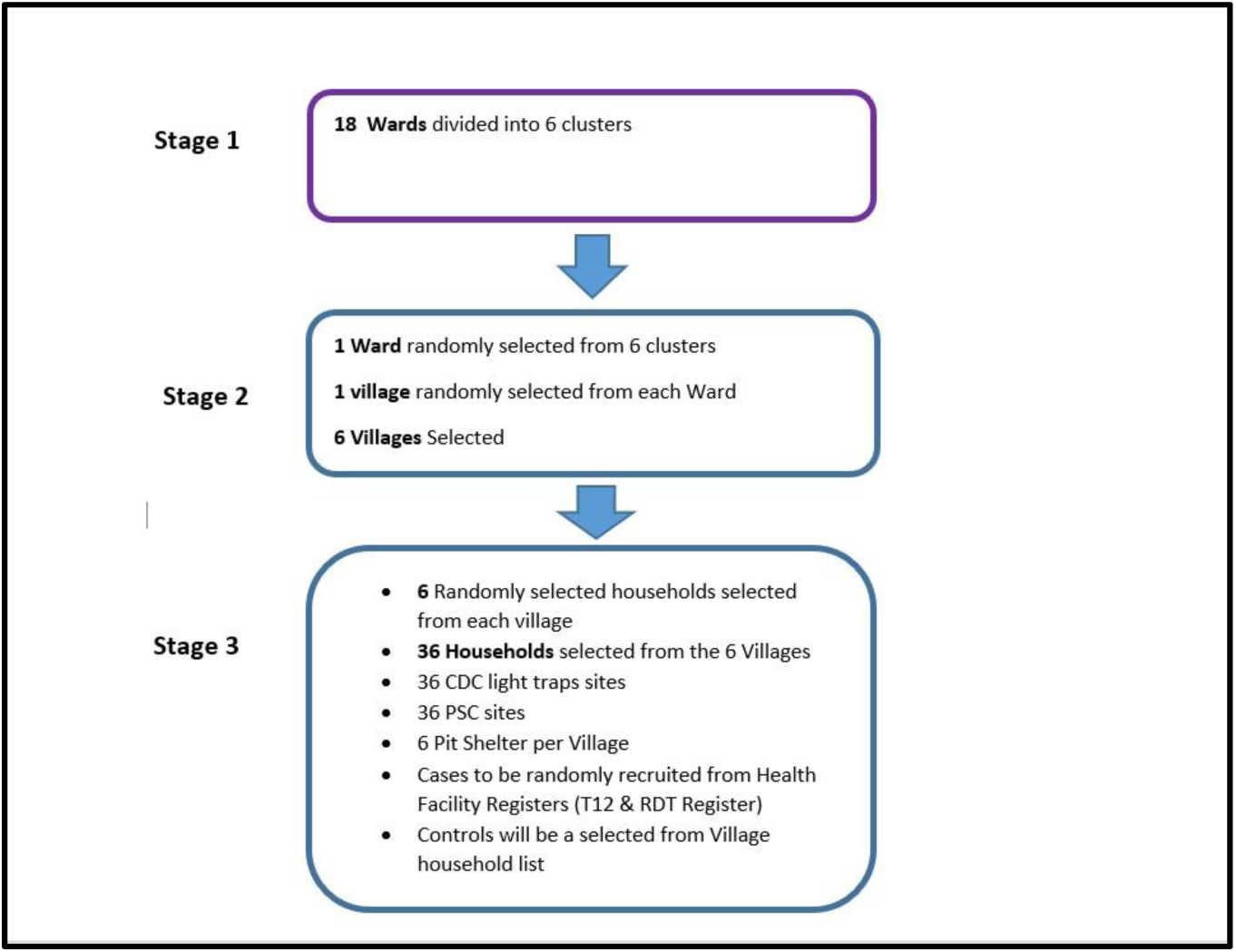
Multi-Stage Sampling Strategy for Malaria in Mudzi Study.

### FGD Questionnaire Design and Data Collection

An FGD guide with eleven (11) guiding questions was utilised to gather data for this study. The FGD guide solicited information on the causes of malaria, signs and symptoms of malaria, treatment sources, challenges in accessing care, perception of current treatment interventions, and preferred malaria control interventions. Perceptions on occupation and social activities related to malaria were also assessed. FGDs were conducted with community members to explore knowledge, practices, attitudes, and perceptions. The FGD had direct and indirect questions (**Supplementary File 2**). Six (6) research assistants (RAs) were recruited to assist with data collection. The RAs were trained for two days on how to administer the form as well as on field ethical practices. Data was collected between July 2021 and August 2021. The FGD guide was prepared in English and translated into *Shona* (the local language). To ensure the consistency of questions, the FGD guide was back-translated into English by an independent person. FGD sessions were held at convenient village meeting places. An audio recorder was used to record all the sessions.

**Supplementary File 2.** Focus Group Discussion Guide: English Version.

|  |  |
| --- | --- |
| 1. What do you think are the most common health problems affecting the community in this Village? | *Probe for all diseases in the community* |
| 2. Do you consider malaria to be a serious disease in this village?<br>*If yes, why do you think so?* |  |
| 3. What do you think causes malaria? | *Probe for beliefs and perceptions* |
| 4. How do we know if someone has contracted malaria? | *Probe for clinical signs and symptoms* |
| 5. Who is mostly affected by malaria in this village? | *Explore all groups, children, mothers, Farmers, gold panners)* |
| 6. When people get sick due to malaria in this village, where do they first seek treatment most of the time? | *Probe for faith healers, traditional healers, private surgeries,*<br>*Probe for home treatment*<br>*Other local providers such as pharmacies, faith healers* |
| 7. What are some of the difficulties you face when trying to seek malaria treatment |  |
| 8. Do you think gardening, farming, artisanal mining, and crossing the border into Mozambique contribute to people getting malaria? |  |
| 9. What is the main source of health information on malaria in this village? |  |
| 10. Do you think spraying homes during the IRS program helps in reducing mosquitoes? |  |
| 11. Which methods of preventing malaria would you prefer in this area | *Probe for any preferred methods of interventions, IRS, treated bed nets, and mosquito repellants.* |

### Public Involvement

The village headman provided local gatekeeper permission to the researchers to conduct the research in the village. Furthermore, the village leadership and business community provided authorization for the use of designated village meeting places for FGD meetings. This ensured that FGDs were conducted at the most convenient places accessible to all participants. The community leadership provided valuable advice on the most appropriate times and days for conducting FGD in each village. Community health workers and community leadership, as well as religious leaders, were critical in promoting participation of the local community in the FGDs.

### Data Collection and Analysis

The recordings were downloaded from the recorder and later transcribed and translated into English for analysis. TF conducted the transcribing, whilst TM verified the transcripts to ensure the translation was accurate and the original meaning was not lost. Open coding was conducted by TF to develop categories and themes that spoke to malaria causes, signs and symptoms, sources of malaria treatment, perception of malaria treatment services, and control. The themes and categories were developed through an iterative process that involved data coding, generation of initial themes, reviewing of developed themes, refining, and naming of themes. JMC and TM reviewed the developed categories. NVIVO Ver 13.1 was used to assist with the analysis and categorization of qualitative data.

### Ethical Consideration

Ethical approval was sought from the University of KwaZulu-Natal Bio-Medical Research Ethics Committee [BREC/00001594/2020] and the Medical Research Council of Zimbabwe [MRCZ/A/2637]. Local permission to conduct the study was obtained from the Provincial Medical Director for Mashonaland East, the District Medical Officer for Mudzi, and the District Administrator. Further permission was sought from the local village headman. Informed written consent was obtained from all the FGD participants. Parental consent was sought for participants below the age of 18 years. Participants were informed of their rights and were free to discontinue participation at any time. Qualitative and Quantitative data collected were kept in secure files.

## Results

### Socio-Demographic Status of Participants

One hundred and forty-eight (148) participants were recruited from six villages, as shown in **Table 1**.

**Table 1.**
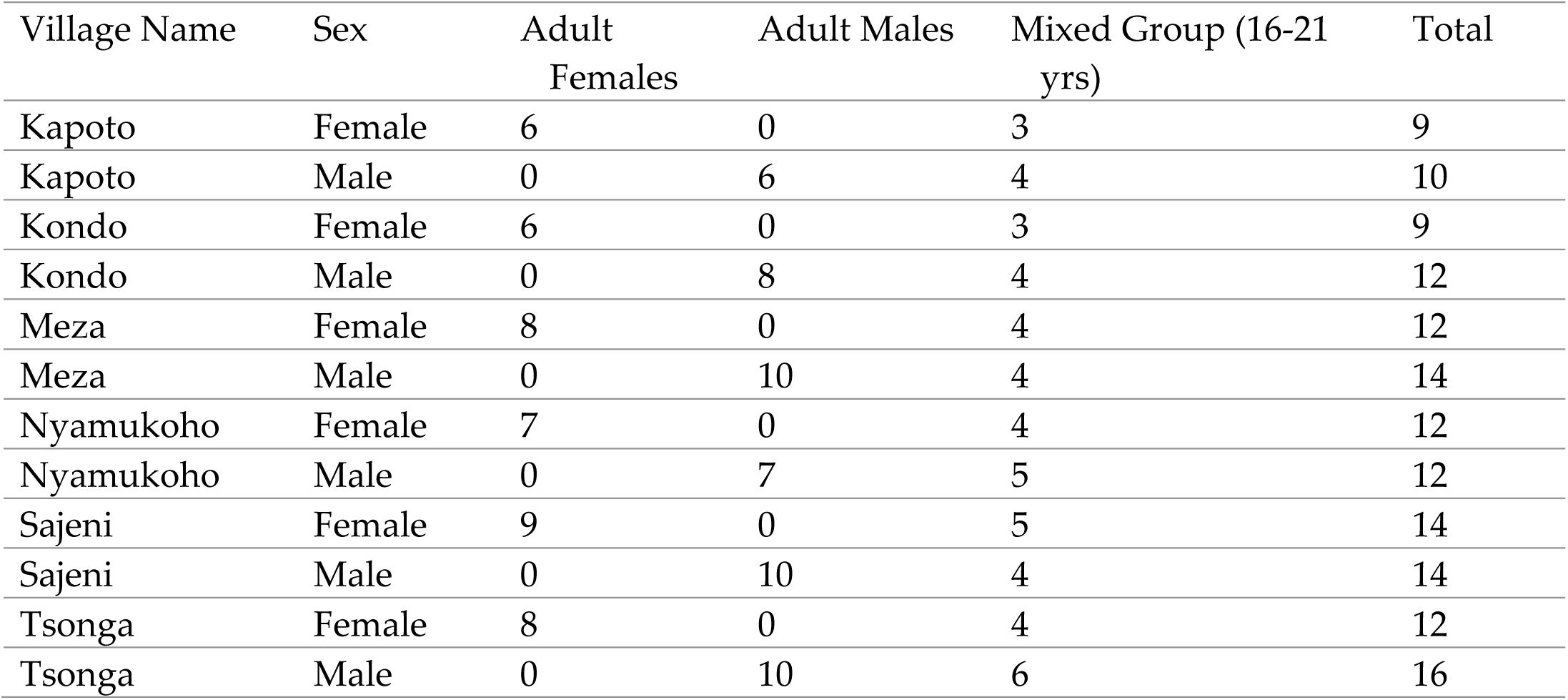
Focus group discussion composition by village and sex.

| Village Name | Sex | Adult Females | Adult Males | Mixed Group (16-21 yrs) | Total |
| --- | --- | --- | --- | --- | --- |
| Kapoto | Female | 6 | 0 | 3 | 9 |
| Kapoto | Male | 0 | 6 | 4 | 10 |
| Kondo | Female | 6 | 0 | 3 | 9 |
| Kondo | Male | 0 | 8 | 4 | 12 |
| Meza | Female | 8 | 0 | 4 | 12 |
| Meza | Male | 0 | 10 | 4 | 14 |
| Nyamukoho | Female | 7 | 0 | 4 | 12 |
| Nyamukoho | Male | 0 | 7 | 5 | 12 |
| Sajeni | Female | 9 | 0 | 5 | 14 |
| Sajeni | Male | 0 | 10 | 4 | 14 |
| Tsonga | Female | 8 | 0 | 4 | 12 |
| Tsonga | Male | 0 | 10 | 6 | 16 |

The majority of the participants were males, 78(52%) aged between 15-19 (31%) and 25-29(21%), **Table 2**. Most of the participants were married (68; 47.2%) and predominantly male. The highest proportion of the participants were unemployed, 102 (69.9%), with only 10 (6.8%) individuals reporting to be formally employed. The majority (138; 94.5%) of the participants had attained secondary school level education.

**Table 2.** Socio-demographic Characteristics of FGD Participants in Six Wards of Mudzi.

| Variable | Female n=(68) | Male (n=78) | n (%) |
| --- | --- | --- | --- |
| **Age** |  |  |  |
| 15-19 | 20 | 26 | 46 (31%) |
| 20-24 | 8 | 5 | 13 (8%) |
| 25-29 | 14 | 18 | 32 (21%) |
| 30-34 | 7 | 8 | 15(10%) |
| 35-39 | 2 | 3 | 5(3%) |
| 40-44 | 4 | 5 | 9(6%) |
| 45-49 | 5 | 3 | 8(5%) |
| 50-54 | 2 | 2 | 4 (2%) |
| 55-59 | 2 | 3 | 5(3%) |
| 60-64 | 3 | 2 | 5(3%) |
| 65-69 | 1 | 1 | 2(1%) |
| 70-74 | 0 | 2 | 2 (1%) |
| **Marital Status** |  |  |  |
| Divorce | 7 (10.3%) | 1 (1.3%) | 8 (5.6%) |
| Married | 29 (42.6%) | 39 (51.3%) | 68 (47.2%) |
| Single | 27 (39.7%) | 36 (47.4%) | 63 (43.8%) |
| Widow | 5 (7.4%) | 0 (0.0%) | 5 (3.5%) |
| Divorce | 7 (10.3%) | 1 (1.3%) | 8 (5.6%) |
| **Employment Status** |  |  |  |
| Employed | 1 (1.5%) | 9 (11.5%) | 10 (6.8%) |
| Self employed | 2 (2.9%) | 13 (16.7%) | 15 (10.3%) |
| Student/pupil | 4 (5.9%) | 9 (11.5%) | 13 (8.9%) |
| Unemployed | 58 (85.3%) | 44 (56.4%) | 102 (69.9%) |
| Village head | 0 (0.0%) | 3 (3.8%) | 3 (2.1%) |
| Community Health Worker | 3 (4.4%) | 0 (0.0%) | 3 (2.1%) |
| **Education** |  |  |  |
| Primary level | 4(5.9) | 4(5.1) | 8(5.5) |
| Secondary | 64(94.1) | 74(94.9) | 138(94.5) |
| Median Age: 27.0 Q1- 18.0- Q3 7.75 IQR: 19.75 |  |  |  |

### Focus Group Discussion Results

#### Malaria is a significant public health threat

The majority of the respondents believed that malaria was a significant threat to livelihood and health, as indicated by many community deaths. They said many young children and mostly pregnant mothers needed treatment. Cases of maternal deaths due to malaria were also reported. Participants in the majority of the FGDs reported that malaria illness at the household level also placed a huge financial burden on the family expenditure, mainly for medication and transport.

> *“When children fall sick, we have to leave our home chores and attend to the sick child. Sometimes, we have to look for transport money to go to the clinic. This is stressful since the majority of us are struggling to make ends meet.”* (K16)-AF

> *“Malaria is a big problem threat here because it costs us as we require money for bus fare and clinic consultation fees”* (K11)-AF

Participants reported that children often miss school lessons due to malaria illness. Participants linked non-school attendance due to malaria as a contributory factor to poor academic performance.

> *“Our children lose learning time when they fall sick, resulting in poor grades. This is usually common during school during the rainy season when a lot of people fall sick”* (K02)-AF

Malaria imposes economic stress on poor families as they often have to look for money for medication and transport to rural health centers.

> *“Malaria is a big problem. We have seen children die and even adult people die. In Meza village, a year ago, a pregnant mother died due to malaria”* (K01). AM

#### Perception of causes of malaria

The majority of respondents were knowledgeable and aware of the cause of malaria. They were aware that mosquitoes were responsible for transmitting malaria. Most FGD participants also correctly identified bites from female mosquitoes as the primary cause of malaria.

> *“I think malaria is caused by mosquitoes which bite people”*(K03)-MG

> *“Malaria is caused by a female mosquito that bites people at night.”* K07 AF

Despite the participants being knowledgeable about the cause of malaria illness, misconceptions were also reported by participants. Participants reported that malaria could be caused by eating unripe local sugar cane *(ipwa),* eating infected fruits, and unhygienic conditions at home.

> *“Eating unripe local sugar cane (ipwa) and infected mangoe fruits causes malaria. My grandmother told me this”* MGF (K11)

> *“Many people have unhygienic habits and practices at their houses. The unhygienic conditions and untidiness cause malaria”* (K08) AM

#### Knowledge of malaria signs and symptoms

Most participants could identify the key malaria signs and symptoms across all FGDs. Fever, joint pains, body weakness, body temperature, chills, and body sweating were the most mentioned key symptoms.

> *“Some people will fall asleep and have general body weakness. Sometimes, we also have rashes and pimples. Some people feel intense headaches and fever. Some people will lose their appetite and will not eat food. Others will feel weak and very cold”* (K07) MGF

#### Malaria vulnerable population groups

Participants had divergent views on who are the most at-risk groups for malaria. Children and pregnant mothers were widely reported as being the most at-risk groups. Older people were also perceived to be at greater risk due to their weak immune systems and aging. Those who were involved in cattle herding, farming as well as those who spent prolonged periods outdoors at night at social business centers and engaged in artisanal gold mining were at risk of contracting malaria.

> *“Everyone here is affected. We all have been affected by malaria. Pregnant women are affected most. Young children are also affected by malaria. Those who do gold panning, attend evening church services and those who frequent beer halls as well as cattle herders get malaria”* (K11)-AM

> *“Women who are pregnant and young children are the most affected. Everyone here gets sick from malaria. Older people are also not spared. Those who work jobs in night jobs and those who drink alcohol at night at social events mostly get malaria. Cattle herders are also not spared.”* (K17)-AF

#### Malaria Treatment Preferences

The majority of the participants indicated that they got treatment services at the local health facilities, but also appreciated treatment services provided by CHW.

> *“Everyone here in the village, when they fall sick, they go to the clinic”* (K17) AF

> *“We get our treatment from the local village health worker as she is closer to us”.* (K16) AM

Participants also mentioned that some community members visit traditional healers and other spiritual healers before they go to the clinic or CHW.

> *“I sometimes go to the faith healer first with my parents, then later go to consult the village health worker”* (K03) AF

> *“We have a large group of African Apostolic faith believers here, and I have seen people go to their prophets first before going to the village health worker”* (K04)-AM

#### Barriers to access to treatment

Participants reported various challenges faced by community members when accessing malaria treatment. The participants reported absenteeism of CHWs from the village. The absenteeism was attributed to social and personal reason personal reasons., The absenteeism affects access to care for the community members, and they usually resort to visiting distant clinics. Participants also mentioned stock-outs of malaria diagnostics and medicines at health facilities and CHWs as a major barrier to treatment. The long distance to the health facility was also cited as a major hurdle to accessing treatment services.

> *“If you fall sick at night, it is difficult to access the CHW. Access general transport or ambulance is impossible in this rural setting at night”* (K03) FMG

> *“Sometimes the clinic is very far and CHWs run out of test kits and malaria medication. This causes us problems as we have to look for money or borrow from relatives or neighbours”* (K14) FMG

#### Perception of Farming, crossing the border, and artisanal activities as drivers for malaria

The majority of participants viewed artisanal mining as a high-risk activity for contracting malaria, mentioning that most of the artisanal gold miners often slept outdoors for longer periods, which exposed them to mosquito bites.

> *“Sleeping in the open exposes one to mosquito bites. Those people will be living in the open, and during the night, they will be exposed to mosquito bites. Artisanal miners create pools at their sites, which breed mosquitoes”* (K04)-AF

Whilst some of the participants viewed artisanal mining as highly risky and associated with malaria illness due to the nature of the work, some participants were of a different opinion. They. viewed artisanal mining as beneficial economically and not a driver of malaria.

> *“Artisanal gold mining brings money, so I don’t see it as a problem which can lead to malaria”* (K13) AF

Participants perceived crossing the border into Mozambique posed a malaria risk, as Mozambique does not implement an IRS program. Travelers were reported to cross the border illegally at night to avoid being arrested by law enforcement agents. This exposes to illegal cross-border to mosquito bites. They also believed that there were more mosquitoes Cuchamano district, where there was no IRS program.

> *“In Mozambique, there is poor hygiene and houses are not sprayed like what is done here. People go for trading there, and often sleep outside exposing themselves to mosquito bites”* (K09) AF

> *“Crossing the border without the protection of mosquito nets and being away from sprayed houses increases the risk of travelers getting malaria”* (K14) AM

Other participants argued that there was no link between crossing the border and malaria.

> *“Crossing the border has no relationship with malaria; it is just for trading”* (K04)-MMG

Some participants believed that gardening provided habitats for mosquito breeding and therefore posed a malaria risk. However, the economic benefits resulting from garden activities were considered to outweigh the risk.

> *“I don’t think gardening has any connection with contracting malaria. We have many people who frequent their gardens, yet they don’t get malaria”* (K02) AM.

> *“In the garden, there are water puddles which are breeding sites. Mosquitoes breed in those puddles and people eventually get exposed to mosquito bites as they work.”* (K07)-AF

#### Source of Health Information for Malaria

Most participants revealed that healthcare and community health workers were the most prominent sources of health education information on malaria. Participants also reported that nurses and EHTs usually gave health education sessions at the health facilities, public gatherings, church gatherings, and funeral gatherings, and immunization outreach activities.

> *“Healthcare workers and CHWs come and teach us here at community gatherings, funerals, and public gatherings”* (K8) AF

> *“The local Environmental Health Technician comes to community gatherings and our households to teach us how to prevent and get treated for malaria”* (K04) AF

Young adults also reported getting information on malaria prevention, signs, and symptoms from school teachers and during health and science lessons.

> *“As pupils, we are taught about malaria by our teachers at school”* (K07) MMG

Participants also reported that television (TV) and radio programs on malaria and health are a prominent source of information. Social media applications were reported by participants as source of information.

> *“Radios on our phones are also a big source of information, and sometimes, to a lesser extent, social media messages on WhatsApp and Facebook”* (K13) FMG

#### Perception of Malaria Control Programs

The majority of participants believed that the IRS chemicals were effective and had contributed to a reduction in malaria cases. Other participants viewed the chemicals as less potent compared to the chemicals used in past IRS programs.

> *“We no longer get a lot of malaria these days compared to the time when we were growing up. I think the chemicals that they use to spray work”* K09 AM

> *“I think these chemicals work; however, I don’t think they are as strong as they used to be years back; they no longer kill cockroaches”* (K05). AF

Participants also reported that the chemicals used in the IRS program sometimes caused body irritations and other discomfort, leading to people sleeping outside.

> *“The chemicals sprayed in our homes cause skin itchiness, and our children and some people will be scratching all the time. This makes it difficult to sleep indoors”* (K16)-AF

Participants were aware of the various methods used at the household level to control mosquito vectors. In addition to home spraying, some participants preferred mosquito nets because they were considered user-friendly and provided better aeration conducive to hot regions like Mudzi than blankets. Some participants preferred the use of mosquito repellents.

> *“We clean our house yard and burn all the rubbish and waste. We cut all the tall grass at our house and close all pits. We make sure all rotten fruits that attract flies are buried”* (K04) AF

> *“We prefer mosquito nets here because when it gets hot, one can easily use them. We would also want mosquito repellent lotions so that when we go to our churches and gardens, we can use them”* (K02) AF

Whilst participants were of the view that the use of IRS should be continued, they also recommended the early onset of household spraying. Other participants believed that whilst nets were ideal, they were prone to abuse by community members as they could be easily repurposed for chicken rearing and illegal fishing activities.

> *“We also want to get mosquito nets. It is hot here, so nets are cooler than blankets and will make sleeping easier. But the nets can be abused as some will use them for their domestic chicken rearing or fishing”* (K06) MMG

## Discussion

Understanding socio-behavioural factors is critical in malaria control. This study assessed the knowledge, perception, attitude, and practices on malaria among community members. The study revealed good knowledge of malaria by participants; however, there were misconceptions about malaria transmission. Health system bottlenecks such as HCW attitude, absenteeism among CHW, malaria medicines stockouts, and long queues at health facilities were considered to be the key hurdles to accessing care.

The majority of the participants were knowledgeable about the causes of malaria and clinical symptoms. Good knowledge of malaria has been observed to be high in endemic settings ^[32]^. This could be attributed to routine and frequent awareness programs carried out in endemic areas. Knowledge of malaria transmission pathways has been linked to health-seeking behavior ^[33]^, whilst a low level of education is a significant risk factor for contracting malaria ^[34]^. Higher levels of education attained by participants may explain the good knowledge revealed in this study^[35][36]^. Education curriculum incorporating disease prevention concepts could therefore play an important role in increasing disease prevention and transmission awareness among the population. Collaborative efforts by the education and health sectors are critical at all levels in the malaria control continuum.

Despite participants being knowledgeable about the causes of malaria, there were also misconceptions about the same. Participants erroneously implicated drinking dirty water, an unhygienic home environment, and eating infected fruits. Misconceptions may result in low-risk perception and a lower likelihood for uptake of preventative measures. Late presentation to care is also likely to result from misconceptions. Misconceptions observed in our study are similar to those noted by Menjetta et al.^[37]^ and Chirebvu et.al in Botswana ^[38]^, where “contact with an ill person,” “poor personal hygiene”, and “staying together, “and “drinking dirty water” were mentioned as causal factors. The corrective action lies in increasing malaria knowledge through continuous malaria health education programs through low-cost, community-appropriate channels ^[39]^.

This study showed that some participants preferred getting malaria treatment from health facilities and CHWs compared to self-medication and faith healers. A high preference for standard care offered through health facilities and CHWs contributes to adequate case management and correct diagnosis, translating to positive treatment outcomes. CHWs complement static health facilities by bringing malaria treatment closer to the community, resulting in reduced transport costs and minimized long queues at health facilities ^[40]^. Evidence from a study in Liberia revealed that CHWs increased access to care in remote rural settings ^[41]^. Improved coverage and presence of CHWs at the community level are often associated with malaria burden reduction ^[42]^. Ensuring uninterrupted access to health facilities and CHWs guarantees access to standard diagnosis and access to effective artemisinin combination therapies (ACT) will likely lead to improved case detection as well as reduced malaria deaths.

Religion plays a critical role in health decisions, as noted by Muyambo et al.; “health decisions are informed by religious and cultural practices” ^[43]^. Studies in Zimbabwe have shown hesitancy among African Apostolic religious groups in seeking orthodox health care ^[44,45]^. Whilst acceptance of orthodox medical services is commendable, reporting at first instance to religious leaders and spiritual healers to obtain “holy waters” and “prayers” before proceeding to an RHC or CHW is a serious concern, as it may lead to delays in seeking care and often compromises management of the disease. Simultaneous interventions are common among religious and spiritual groups ^[43]^ and may affect treatment adherence. Religious practices and beliefs are significant determinants of malaria, as shown in studies in Ghana and the Democratic Republic of Congo ^[46,47]^. It is critical to create community-based programs that are sensitive to prevailing religious practices and inclinations in areas where there is strong religious participation.

HCW and CHW negative attitudes, long queues, lack of transport, and inadequate CHWs are serious barriers to malaria treatment and care. The behavior of health care providers and the availability of essential commodities, infrastructure, and services affect health user satisfaction ^[48]^. The negative health worker attitude and stock-outs may lead to reduced confidence in standard care at health facilities and may influence delays in seeking care ^[49]^. Delays in seeking standard care have been associated with the development of complicated and severe malaria cases ^[50]^. Similarly to our findings, Chuma et al. ^[51]^ in Kenya identified “availability” and “provider-patient relationship” as key barriers. In the same study, HCWs were reported to be “inconsiderate” and “uncaring”^[51]^. There is a need to improve health care provider quality of service through in-service training in patient-centered quality improvement programs ^[52]^. A rural health facility is manned by two (2) nurses who have to deliver a wide array of services, ranging from maternal and child health services, general outpatient care, immunization services, as well as malaria testing and treatment. This tends to increase the work overload, and hence increasing staffing through the recruitment of additional CHWs and HCWs reduces staff burnout and patient waiting times. Improving the availability of healthcare workers has been shown to decrease complicated cases and deaths ^[53]^.

The mixed views on the risks posed by illegal artisanal gold mining and cross-border movement reveal the dilemma of communities whose livelihoods depend on the same activities for survival against the backdrop of existing high unemployment. The two activities have been significantly associated with the occurrence of malaria elsewhere ^[54]^ yet they are vital to survival in communities such as Mudzi. The presence of a porous border with no fence also facilitates human movement between Mudzi and Mozambique. Unmanned points of entry have also been shown to be used by border communities and illegal immigrants who lack proper documentation and may also be involved in smuggling activities ^[55]^. Migrating populations often lack access to malaria protective measures^[55]^ and increase the risk of contracting malaria. The combined effect of artisanal mining along border regions and human cross-border movement significantly drives persistent malaria transmission in other regions ^[54,56,57]^. Countries with endemic malaria in border regions should upscale community engagement programs, increasing border control and enhancing cross-border malaria collaborative control mechanisms^[55,57]^. The creation of community action groups could be pivotal in reducing risk among cross-border and artisanal gold miners because they can be very effective in improving risk perception, knowledge, and preventative behavior ^[58–60]^.

The IRS has been the principal core vector control intervention responsible for reducing the malaria burden globally in the past decades. However, participants believed that the chemicals used in IRS programs were not efficacious. This may indicate an ineffective IRS awareness and sensitization program, as the various chemical actions, reasons for rotation, and effectiveness should be explained fully to program recipients. Reported side effects due to IRS chemicals may create low uptake and hesitancy. Negative effects such as headaches, respiratory disorders, discoloring of inner household walls, and unpleasant odours have been reported in other studies as factors contributing to non-acceptance of IRS ^[61,62]^. Effective communication of the IRS side effects and mitigatory measures needs strengthening. Evidence has shown additional protection by adding LLINS to IRS, but that creates an additional cost which many low-income countries cannot afford ^[63,64]^. The preference for complementary LLINS/ITNs by communities was also reported in a similar study in Mozambique ^[65]^. However, innovative ways to deliver such large-scale dual vector control interventions at a low cost are yet to be found.

## Limitations

Our findings and conclusions are based on unverified self-reported views from participants. The selection of knowledge-rich individuals by the CHW introduced bias, as those who volunteered may have already been active, prominent, and visible members of the community. Other less willing, hesitant individuals who could have been knowledgeable may have been excluded, thus depriving the FGD of diverse views. As a strength, the use of separate groups for females, males, and young adults ensured all community groups expressed themselves freely without being encumbered by social norms. Furthermore, our study represents a first attempt to document knowledge, attitudes, and practices that could likely explain human behaviour drivers to persistent malaria transmission in the district.

## Conclusion

Our study revealed complex interrelated socio-demographic, behavioural, economic, and health system factors that may explain the sustained, persistent malaria transmission in Mudzi district. Targeted interventions for religious groups and artisanal gold miners should be developed to reduce misconceptions and nighttime outdoor movement.

Strengthening client-centered care approaches among HCWs and CHWs may assist in reducing negative perceptions among health care users. Eliminating malaria in highly endemic regions with persistent transmission, such as Mudzi, will require significant investment by health authorities and the private sector in expanding human resources for health, malaria diagnostics and medicines, community treatment, and adequately equipping CHWs.

## Data Availability

All data produced in the present work are contained in the manuscript

## Author Contributors

TF conceived and designed the study. MJC and TM contributed to the study design.TF and TM conducted the data analysis. TF drafted the manuscript. MJC and TM critically reviewed the manuscript. All authors read and approved the final version.

## Ethics approval

Ethical approval was sought from the University of KwaZulu-Natal Bio-Medical Research Ethics Committee [BREC/00001594/2020] and the Medical Research Council of Zimbabwe [MRCZ/A/2637].

## Data sharing statement

All data relevant to the study are included in the article.

